# The use of compassionate Ivermectin in the management of symptomatic outpatients and hospitalized patients with clinical diagnosis of COVID-19 at the Medical Center Bournigal and the Medical Center Punta Cana, Rescue Group, Dominican Republic, from may 1 to august 10, 2020

**DOI:** 10.1101/2020.10.29.20222505

**Authors:** José Morgenstern, José N. Redondo, Albida De León, Juan Manuel Canela, Nelson Torres, Johnny Tavares, Miguelina Minaya, Óscar López, Ana María Plácido, Ana Castillo, Rafael Peña Cruz, Yudelka Merette, Marlenin Toribio, Juan Asmir Francisco, Santiago Roca

## Abstract

No antiviral has been shown to reduce mortality in SARS-COV-2 patients to date. In the present observational and retrospective report, 3,099 patients with a definitive or highly probable diagnosis of infection due to COVID-19 were evaluated between May 1st to August 10th, 2020, at Centro Medico Bournigal (CMBO) and Centro Medico Punta Cana (CMPC), and all received compassionate treatment with Ivermectin. A total of 2,706 (87.3%) were discharged for outpatient treatment, all with mild severity of the infection. In 2,688 (99.33%) with outpatient treatment, the disease did not progress to warrant further hospitalization and there were no deaths. In 16 (0.59%) with outpatient treatment, it was necessary their subsequent hospitalization to a room without any death. In 2 (0.08%) with outpatient treatment, it was necessary their admission to the Intensive Care Unit (ICU) and 1 (0.04%) patient died. There were 411 (13.3%) patients hospitalized, being admitted at a COVID-19 room with a moderate disease 300 (9.7%) patients of which 3 (1%) died; and with a severe to critical disease were hospitalized in the ICU 111 (3.6%), 34 (30.6%) of whom died. The mortality percentage of patients admitted to the ICU of 30.6%, is similar with the percentage found in the literature of 30.9%. Total mortality was 37 (1.2%) patients, which is much lower than that reported in world statistics, which are around 3%.

---

Coronaviruses are a group of diverse, large RNA viruses that have been known since the 1960s. Most live in animals, but there are 7 of them that are recognized as capable of infecting humans. Two coronaviruses have produced outbreaks of severe lower respiratory infections in people: Severe Acute Respiratory Syndrome due to Coronavirus 1 (SARS-CoV-1), which emerged in southern China in 2002, and Middle East Respiratory Syndrome (MERS)), which appeared in the Middle East in 2012 and which, unlike SARS, still continues with sporadic cases in that geographical region.

In December 2019, a new coronavirus (SARS-COV-2) was identified in Wuhan, China, and declared a pandemic in March 2020 by the World Health Organization (WHO). It produces a disease known as Severe Acute Respiratory Syndrome Coronavirus 2 (SARS-CoV-2). (1,2)

At Monash University in Australia, Leon Caly and a group of collaborators infected Vero-h SLAM cells with SARS-COV-2 in vitro and added ivermectin only once within 2 hours of infecting the cells. After adding ivermectin, 48 hours later there was a 99.8% reduction in viral RNA compared to another control sample to which Ivermectin was not added. The 100% effective concentration of Ivermectin in this study was 5 microMoles and the 50% effective concentration of Ivermectin was 2 microMoles (minimum viral inhibitory concentration). (3,4)

The Ethics Committee and the Corporate Medical Directorate of Grupo Rescue, in their weekly meeting, based on the absence of availability of Hydroxychloroquine in the 2nd and 3rd week of April 2020, when they received a high number of critical cases of patients infected with SARS-COV-2 in the Emergency Service (ER) of the Bournigal Medical Center (CMBO), Rescue Group in Puerto Plata, they decided to modify the institutional management guide to include Ivermectin based on the in vitro study of the Monash University(3) in conjunction with Azithromycin for both inpatient and outpatient treatment. The present study is a report of the observed results.

## Material and Methods

### Objectives

1. To show that Ivermectin reduces mortality from COVID 19 infection.
2. To demonstrate that in patients in phase I of the disease, Ivermectin slows the progression of the disease, decreases the number of hospitalizations and decreases the number of deaths.

## Design

### Retrospective observational study

From May 1 to August 10, 2020, the drug Ivermectin was indicated as a compassionate treatment to all patients admitted in the ER of the Bournigal Medical Center (CMBO) and the Punta Cana Medical Center (CMPC), with the definitive diagnosis of COVID -19 due to chain reaction of real-time polymerase transcription (rt-PCR), or with a highly probable diagnosis due to the presence of at least 1 of the major criteria of its protocol and at least 3 of its minor criteria (Table 1).

**Table 1:**
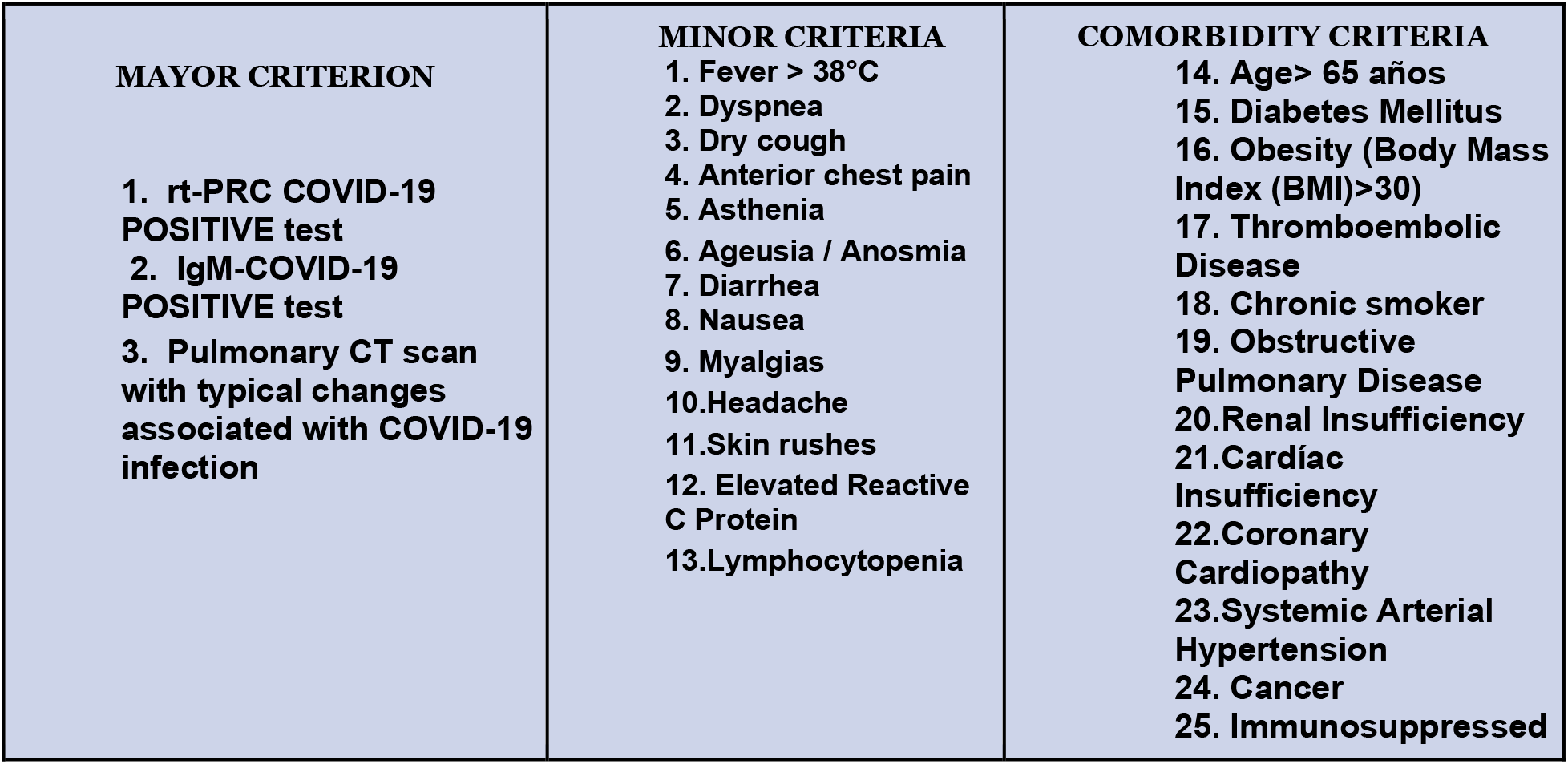
Major, Minor and Comorbidity criteria for the diagnostic of infection of COVID-19

Pregnant women, breast feeding women, children under 2 years or under 15 kg, patients allergic to the drug or those who are anticoagulated with coumarins are excluded for the administration of Ivermectin. In addition, patients who, once informed of the pros and cons of the use of the drug for the treatment of COVID-19, refused its use and refused to sign the informed consent designed for this purpose, were also excluded from treatment with Ivermectin.

All patients received in the ER undergo: rt-PCR or Immunoglobulin (Ig) M by the rapid qualitative test for COVID 19 depending on availability, pulmonary Computed Tomography (CT) scan and laboratory tests: complete blood count, quantified reactive C protein, glycemia, creatinine, transaminases, lactate dehydrogenase (LDH), ferritin, D-dimer, swab for influenza A and B, and a standard 12-lead electrocardiogram.

A scale of severity was established to decide the patients to be treated outpatient or hospitalization, namely:

- Grade 1 (mild): patients with arterial oxygen saturation greater than 93% room air, heart rate less than 125 / minute, respiratory rate less than 24 / minute, no decompensated organs and no deterioration of comorbidities (Table 1).
- Grade 2 (moderate): Sustained arterial oxygen saturation (3 minutes) equal to or less than 93% room air, dyspnea, respiratory rate greater than 24 / minute, heart rate greater than 125 / minute.
- Grade 3 (severe): sustained arterial oxygen saturation (3 minutes) equal to or less than 80% ambient air, arterial hypotension.
- Grade 4 (critical): Patients Grade 3 who do not respond to support measures and require mechanical ventilation, administration of vasopressors or hemodialysis.

Grade 1 patients were treated on an outpatient basis and followed up by our medical staff, with instructions to return immediately to the ER if they had disease progression. Grade 2 patients were hospitalized in isolation rooms in the COVID-19 area; and grade 3 and 4 patients were hospitalized in the Intensive Care Unit (ICU) in the COVID-19 area.

The outpatients were administered Ivermectin at 0.4mg / kg, orally (PO) in a single dose in the ER and Azithromycin 500mg PO per day for 5 days, with follow-up of the outpatients.

Hospitalized patients were administered Ivermectin PO at 0.3mg / kg, days 1 and 2, and the dose was repeated on days 6 and 7. They were given Azithromycin 500mg PO daily, for 7 days.

If they presented a D-dimer greater than 1,000 ng / ml or an increase of 50% from the initial value, they were started with Enoxaparin at 1mg / kg subcutaneously, every 12 hours. Patients with an elevated D-dimer, but less than 1,000 ng / ml received Enoxaparin at 1mg / kg subcutaneously, every 24 hours. Enoxaparin was not administered if there was thrombocytopenia less than 50,000. (5)

Patients who required oxygen received Dexamethasone at 0.1mg / kg PO per day, maximum 10mg per day, for 10 days (or Methylprednisolone at an equivalent dose). (6)

Critically ill patients with suspected Cytokine Cascade Syndrome, due to radiological progression and involvement of more than 50% of the lung fields that require high oxygen flow or mechanical ventilation or Bilevel Positive Airway Pressure (BIPAP), with increased inflammatory reactants as reactive C protein, D-dimer greater than 1,000 ng / ml and ferritin greater than 500 micrograms / L, they were administered Interleukin 6 Blockers, of the Tocilizumab type, 400mg intravenously and the dose was repeated after 24 hours if there was no clinical improvement. If Tocilizumab was not available, they were administered Methotrexate, a Jak /Stat blocker at 1mg / Kg on days 1, 2 and 3 intramuscularly or intravenously. Immunosuppressed patients, with mild to moderate renal insufficiency, ages older than 70 years, were administered a lower dose of 0.5mg / Kg on days 1,2 and 3 via intramuscular or intravenous. Methotrexate was not administered if the creatinine clearance was less than 30mg / dl. At 48 hours after the last dose of Methotrexate, folic acid 15mg daily was started PO for 7 days. (7,8,9).

## Findings and Results

A total of 3,099 patients were evaluated in the ER of the CMBO and CMPC from May 1 to August 10, 2020 and treated with Ivermectin.

Initially, 2,706 patients (87.3%) received treatment on an outpatient basis, of which 2,688 (99.33%) did not progress the disease, so they did not merit new admission to the ER and subsequent hospitalization and there were no deaths. Of the patients treated as outpatients, 16 (0.59%) subsequently merited hospitalization in the COVID-19 area room with 0 (0%) deaths and 2 of them (0.08%) required hospitalization in the ICU, of which 1 died (0.04%). (Table 2).

**Table 2:**
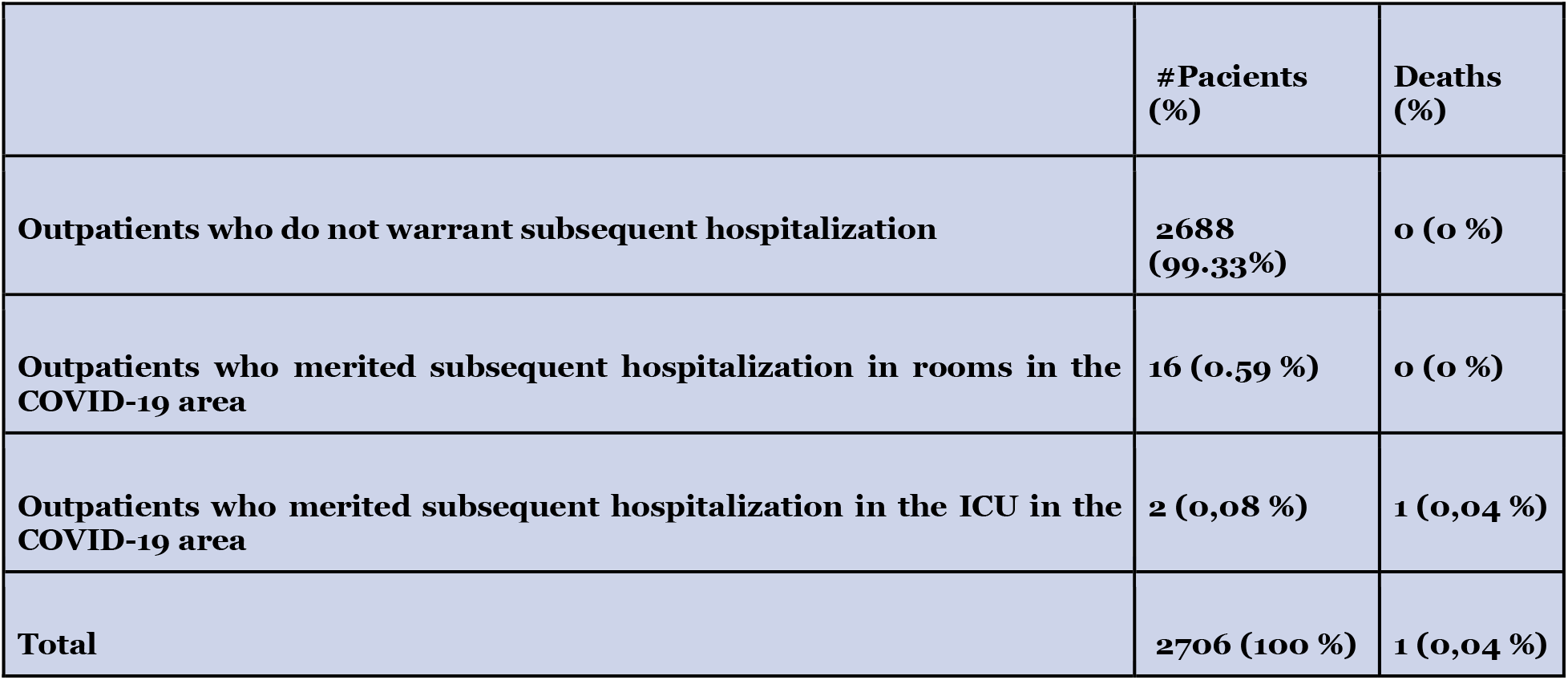
Patients discharged from the Medical Center Bournigal and Medical Center Punta Cana Emergencies for outpatient treatment

In total, 411 patients (13.3%) were hospitalized, including patients initially treated on an outpatient basis and later merited hospitalization, of which 300 patients (73%) were admitted to rooms in the COVID-19 area, representing 9.7% of all patients admitted in the ER. On the other hand, in the COVID-19 ICU, 111 patients (27%) were hospitalized, representing, 3.6% of the cases originally treated in ER. (Table 3 and Table 4)

**Table 3.**
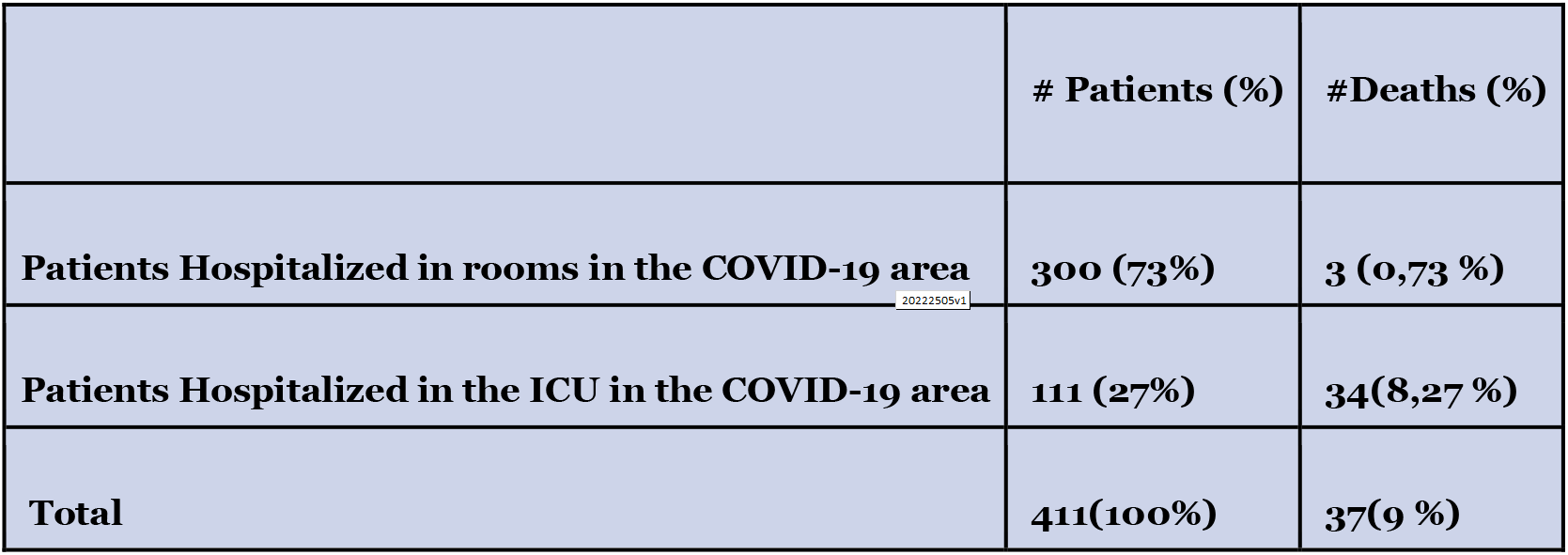
Patients Hospitalized at the Medical Center Bournigal and Medical Center Punta Cana

**Tabla 4:** Medical Center Bournigal and Medical Center Punta Cana

|  | #Patients (%) | #Deaths (%) | % Deaths in each subgroup |
| --- | --- | --- | --- |
| **Outpatients who do not warrant subsequent hospitalization** | **2688 (86,7%)** | **0 (0 %)** | **0 %** |
| **Patients Hospitalized in rooms in the COVID-19 area** | **300 (9,7 %)** | **3 (0,1 %)** | **1 %** |
| **Patients Hospitalized in the ICU in the COVID-19 area** | **111 (3,6 %)** | **34 (1.1 %)** | **30,6 %** |
| **Total** | **3099(100%)** | **37 (1,2 %)** | **1,2 %** |

Of the total number of hospitalized patients, 37 deaths (9%) were reported. Three (3) of them (8%) occurred in the regular isolation area, while 34 (92%) occurred in the COVID-19 ICU (table 4). The total mortality of the patients initially evaluated in the ER was 37 (1.2%). (Table 3 and Table 4)

From the patients admitted to the ICU, 30.6% with degrees of severity 3 and 4 of the disease, died. (Table 4)

The average between the onset of symptoms and the ER visit in outpatient treated patients, was 3.6 days, while in patients hospitalized in a COVID-19 isolation room it was 6.9 days, and in patients hospitalized in ICU-COVID-19 of 7.8 days.

The distribution by gender of all patients admitted to the ER was as follows: 50.02% male and 49.98% female. While the distribution in hospitalized patients was: 63.5% male and 36.5% female. (Table 5)

**Tabla 5:**
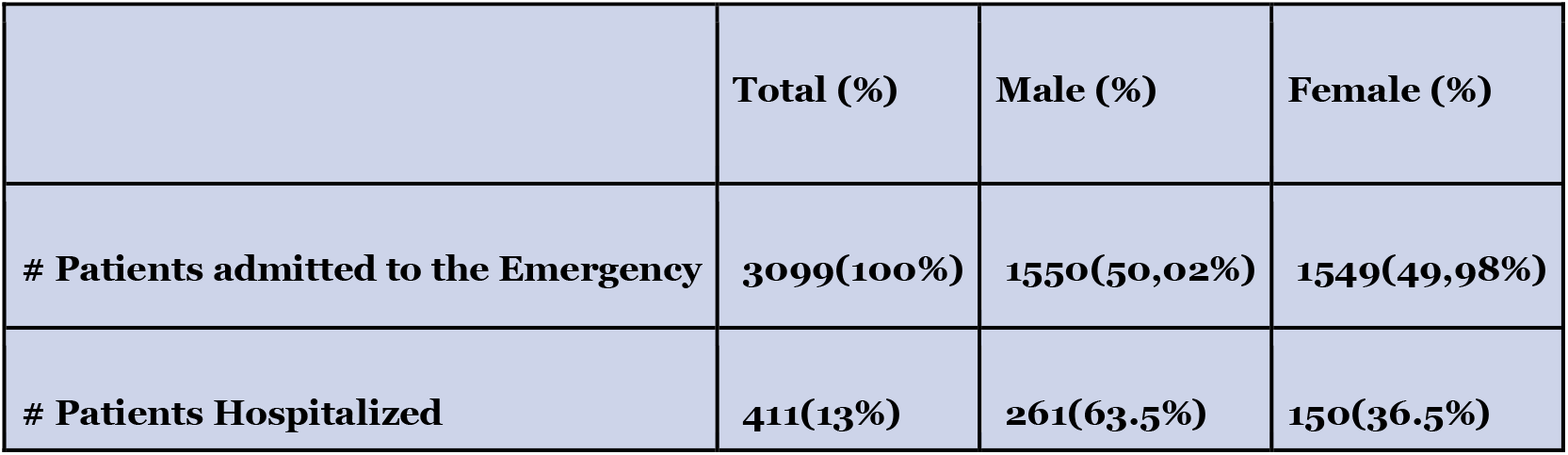
Gender Distribution

|  | Total (%) | Male (%) | Female (%) |
| --- | --- | --- | --- |
| **# Patients admitted to the Emergency** | **3099(100%)** | **1550(50,02%)** | **1549(49,98%)** |
| **# Patients Hospitalized** | **411(13%)** | **261(63.5%)** | **150(36.5%)** |

The average age of the patients hospitalized in rooms of the Covid-19 isolation area was 52 years; while in ICU patients it was 58 years.

COVID-19 rt-PCR tests were performed only on a total of 685 patients, equivalent to 22% of all patients admitted in the ER with a clinical diagnosis of Covid-19 according to the institutional management guide; of these, 360 (52.5%) were reported positives (DETECTED) and 325 (47.5%) negatives (NOT DETECTED). (Table 6)

**Tabla 6:** rt-PCR COVID-19 test

|  | **Positive (%)** | **Negative (%)** | **Total (%)** |
| --- | --- | --- | --- |
| **rt-PCR COVID-19 test** | **360(52.5%)** | **325(47.5%)** | **685(100%)** |

On the other hand, IgM COVID-19 tests were performed on a total of 2,041 (65%) patients, of which 413 (20%) were positives and 1,628 (80%) negatives. (Table 7)

**Table 7:** IgM COVID-19 test

|  | **Positive (%)** | **Negative (%)** | **Total (%)** |
| --- | --- | --- | --- |
| **IgM COVID-19 test** | **413(20%)** | **1628(80%)** | **2041(100%)** |

## Discussion

Ivermectin inhibits the import of SARS-COV-2 nucleocapsid proteins into the nucleus of the infected cell, by blocking Importin (IMP) alpha / Beta 1. These viral nucleocapsid proteins block the antiviral response produced in the nucleus of the infected cell. (10)

Ivermectin is a powerful helicase inhibitor, a very important enzyme, since its function is to unwind the viral Ribonucleic Acid (RNA). Blocking helicase prevents viral replication. (11)

SARS-COV-2 can enter cells in two ways: via endosomes activated by Cathepsin L, or by fusion with the membrane activated by Transmembrane Serine Protease (TMPRSS2). In both situations, the Spike (S1 and S2) Protein binds to the Angiotensin Converting Enzyme 2 (ACE2) receptor. The entry of the virus through the membrane fusion mechanism is the most efficient since it is less likely to trigger an antiviral immune response and is more efficient in producing the viral replication. (12,13)

Ivermectin prevents the binding of SARS-COV-2 S Protein, with the ACE2 receptor and with the CD147 transmembrane receptor. The binding of SARS-COV-2 S Protein with the ACE2 receptor is essential for the virus to enter cells at the time of its replication (to obtain this effect, the total administered dose of Ivermectin must be 2mg / Kg). The CD147 receptor is abundantly distributed in red blood cells, which when joining the SARS-COV-2 S Protein, produces an accumulation of red blood cells at the endothelial layer together with white blood cells and platelets, which causes vascular obstructive phenomena. (14)

On the other hand, Ivermectin can bind to 3CL protease and HR2 protein, blocking viral replication. (15)

It is very true and evident that there are other factors to consider when the patient receives a drug, and that surely play an important role in the clinical response. Namely:

- Humoral immune response of the host against SARS-COV-2 by B lymphocytes and cellular by T lymphocytes, NK lymphocytes, macrophages, dendritic cells. Response of memory lymphocytes acquired by previous infections with other coronaviruses.
- The concomitant administration of Azithromycin, which has a recognized anti-inflammatory, immunomodulatory, and even antiviral effect (in vitro, its effective concentration of 50% antiviral SARS-COV-2 is 2.12 microMoles) (16). Reduces the secretion of the Tumoral Necrosis Factor (TNF) which results in its anti-inflammatory effect. In addition, it inhibits mTORC1 (Mammalian Serine / Threonine Protein Kinase). The latter is essential for COVID-19 to be able to replicate, to be able to infect other people and to disguise itself from the immune system (SARS-COV-2 is not recognizable by the immune system the first 4 days after infecting the patient). In addition, mTORC1 causes a delay in the response of the immune regulatory system that makes it easier for patients infected by COVID-19, especially older adults or patients with pre-existing medical conditions, to present an excessive immune response. (17,18,19,20)

At the time of this presentation of clinical data, there is no controlled, randomized, double-blind, and peer reviewed study in which an antimicrobial has shown, without any doubt, a decrease in mortality of hospitalized patients due to COVID-19 infection.

Remdesivir showed a decrease in the hospitalization time of the patients who received it, but it did not show a significant decrease in mortality compared to the control group. (6,21)

Studies conducted in France using Hydroxychloroquine alone or in combination with Azithromycin also did not show a decrease in mortality (22,23,24). A study reported in China when Lopinavir / Ritonavir was administered, versus placebo did not show a decrease in mortality (25).

The Oxford University Recovery study discontinued the Hydroxychloroquine and Lopinavir / Ritonavir treatment arms due to their ineffectiveness. Its greatest contribution was that it reported a significant decrease in mortality of 17% in oxygen-dependent patients and 33% decrease mortality in patients in mechanical ventilation, when receiving dexamethasone. (6)

A study in Broward / Fort Lauderdale, USA compared 173 hospitalized patients who received Ivermectin versus 107 hospitalized patients who did not receive Ivermectin. The mortality of the group that received Ivermectin was 15% versus 25.2% of the group without Ivermectin. Mortality in hospitalized patients with severe lung disease in the Ivermectin group was 38% versus 80.7% in the no Ivermectin group. (26)

In a study carried out in 63 outpatients in Peru with COVID 19, all patients received Ivermectin and after 48 hours, most symptoms disappeared. In no case was there progression of the disease that warranted hospitalization. (27)

A group from Bangladesh studied 181 outpatients. One group was treated with Ivermectin + Doxycycline versus another with Hydroxychloroquine + Azithromycin. In the Ivermectin group, 100% of the patients recovered without disease progression in an average of 5.93 days and a negative rt-PCR test in 8.93 days. In the Hydroxychloroquine group, 96.3% of the patients recovered without disease progression on average in 6.99 days and a negative rt-PCR in 9.33 days. In 2 patients of the Hydroxychloroquine group, 3.7%, there was progression of the disease and merited their hospitalization. (28)

At the time of reporting this study, there are 40 investigations in progress, registered in the Clinicaltrial.gov portal, in the preprint stage, some of them with preliminary results that seem to support the findings of our scientific observation.

Because it is a new disease, with a significant capacity for contagion between people and a high mortality rate in severe cases, 4 different challenges have been posed:

1. Definition of the clinical symptoms and signs that allow us to recognize the disease with a high degree of efficiency. (29)
2. Laboratory tests and images that allow us to diagnose the disease, identify in which phase of evolution it is, and the degree of severity and prognosis of it. (30,31,32) These phases are
  a. Phase I in which viremia occurs, and is recognized by the appearance of symptoms, on average after the 5-day incubation period of the virus in the host.
  b. Phase II or Pulmonary Phase, which is recognized by the appearance of pneumonia, on average 8 days after the onset of symptoms.
  c. Phase III or Hyperinflammatory Phase, which is recognized by the Cytokine Cascade, Macrophage Activation Syndrome and a vascular thrombotic response. This severe phase appears between days 10 and 19 after the onset of symptoms. (5,7,8)
3. To have different sensitive and specific rt-PCR tests and any other method of direct detection of the SARS-COV-2 virus (currently under development). To have reliable tests IgM and IgG antibodies, sensitive and specific, that allows us to complement the diagnosis of the disease and even recognize a protective post-infection immunity to new COVID-19 infections. (33.34)
4. To have an accessible antiviral treatment, administered orally, safe and inexpensive, that not only allows us to treat the infection in hospitalized patients and reduce their mortality, but that is also effective in outpatients and thus will prevent the progression of the disease, in order not to saturate the hospital services, reduce the duration of the disease and shorten the time of transmission of the virus from the patient to healthy people. Also, to have adequate treatments for complications that may arise during the progression of the disease.

Ivermectin is a drug that has been used extensively and massively for more than 30 years, in patients in Africa and many other countries, for the treatment of blindness causing onchocerciasis and strongyloidiasis, among many other endoparasites and ectoparasites. Very few side effects are known, it is a safe drug (the 50% lethal dose is 29.5mg / Kg), inexpensive, administered orally and with little interaction with other known drugs, except that it enhances the action of warfarin. (10, 35, 36, 37)

It is very important to be able to treat viremia in all 3 phases of the disease. In phase I in which there is more viral replication, the viral load is still lower than in the following phases, so it is probably possible to control the disease with lower doses of the antiviral. In phases II and III the viral load is high. It is already known that the average viral load in patients who survive is 158,498 copies per ml, while the average viral load in patients who die is 2,511,886 copies per ml. For each logarithm that viral load increases, mortality increases by 7%. (38)

A committee made up of specialist physicians, biostatistical researchers and other collaborators from the Rescue Group member hospitals, made decisions based on the available information, in order to face these 4 challenges mentioned above. The committee meets weekly since March 2020, to develop and update the guidelines for the management of COVID-19, an unknown disease and with new information daily.

The committee decided to create a Table with Major, Minor and Comorbidity criteria, which would allow us to make the diagnosis of COVID-19 infection. The presence of 1 major criterion and 3 minor criteria is considered sufficient to make the clinical diagnosis of COVID-19 infection.

In addition to the positive rt-PCR test, two other tests were included in the Major criteria: positive IgM antibodies to COVID-19 and a pulmonary CT scan with typical radiological changes suggestive of COVID-19 infection. Several reasons compel us to not rely solely on the rt-PCR, among which stand:

- Little availability of it in a large part of the pandemic or the delay in obtaining the result;
- Its sensitivity is 67% according to the literature reviewed;
- It depends on an adequate taking of the nasopharyngeal sample, on the day of the disease on which the sample is taken, being positive from 2 days before the appearance of symptoms. The sensitivity decreases progressively after the 10th day, which is precisely when the most severe conditions are seen;
- Positivity depends on a minimum number of viral copies capable of detecting the test, on the preservation of the cold chain in handling the sample until it is read in the laboratory, and on its processing and adequate reading. (33)

IgM-COVID-19 becomes positive 8 days after onset of symptoms and peaks at 12 days. Its sensitivity is as poor as 64.8%. (34)

Pulmonary CT scan shows very suggestive, although not pathognomonic, typical changes associated with COVID-19 infection, which include the early presence of a ground glass image, peripherally or subpleural located, with generally bilateral involvement, as a finding initial, which has been found present in 97.6% of patients. In addition, consolidated bilateral pneumonic type, present in 72.7% of the patients and crazy pavement usually of subpleural or peripheral distribution, often with central preservation, present in 70% of patients, between days 8 and 14 of the appearance of symptoms. Diagnostic changes of COVID-19 Pneumonia in CT scan tend to last a long time to return to normal, even after the disappearance of symptoms (31).

Symptoms, signs and laboratory results were included in Minor Criteria, among which are fever greater than 38 °C, dyspnea, dry cough, chest pain, asthenia, ageusia, anosmia, diarrhea, nausea, myalgias, headache, skin rashes, elevated reactive C protein, lymphocytopenia. (30,32) (Table 1)

Conclusions of the therapeutic retrospective observation in patients initially seen in the emergency room and followed up on an outpatient basis or hospitalized:

1. In 99.3% of the outpatients who were treated with Ivermectin, the establishment of treatment was effective in the early stages of the disease (on average 3.6 days from the onset of symptoms), since the infection did not progress. They did not merit subsequent hospitalization and had no deaths. Only 0.67% of the outpatients who were treated with Ivermectin, did the disease progress and merit hospitalization, with 1 death (0.04%).
2. All hospitalized patients received Ivermectin and it was observed that the mortality of the severe to critical patients hospitalized in the ICU, was similar to that reported in the international literature of 30.9% versus 30.6% in the patients of the present study (39).
3. Total mortality adding up outpatients and hospitalized patients treated with Ivermectin, was 1.2%, well below the average 3% reported in most series and overall mortality worldwide. (40) It is important to recognize that our Lethality Rate is compounded by the number of fatalities over the number of “clearly sick” patients who attended to our ER, for proper diagnosis and treatment.
4. No severe side effects were observed with the administration of Ivermectin. The only effects reported by some patients were mild nausea, heartburn or mild diarrhea, which in no case merited the suspension of treatment. No fatal or severe arrhythmias associated with the use of Azithromycin were observed.
5. In patients who came to the ER for a COVID-19 infection, by clinic, by laboratory and with typical changes in the pulmonary CT scan, and who could have a sample taken for rt-PCR, only in 52.5 % of the cases the test was reported as positive. This result is below that observed in the literature of 67% and warrants a review in the methodology of taking, handling and processing the sample.
6. Those who underwent IgM COVID-19 were only positive in 20% of the patients, also well below expectations of 64.8%, which may be related to the day of the disease that the sample is taken counting from the onset of symptoms or the sensitivity of the test itself.
7. These findings reinforce the claim that the diagnosis of this disease cannot be exclusive to the COVID-19 rt-PCR test. The clinic, laboratory and imaging (especially pulmonary CT scan) are essential to make the diagnosis. For this reason, we recommend the use of the Table of Major and Minor Criteria to establish the diagnosis of COVID-19 infection. (Table 1)
8. The distribution by gender of the patients who attended the ER was the same in the relation of men and women. However, in hospitalized patients, the proportion of men was 63.5% versus women 36.5%. This coincides with the findings in other studies and it has been proposed that it could be related to the presence of higher levels of androgens in man, which stimulates the production of TMPRSES2, which is a protease that facilitates the entry of the virus into the cell once the spike (Protein S) binds to the ACE2 receptor. (12,13,41)

## FINAL NOTE

These results obtained are retrospective and observational. A prospective, randomized and double-blind comparative study is required with the use of ivermectin, to be able to confirm without doubts and by analysis of pairs these findings in outpatients and in hospitalized patients.

## Data Availability

The data was collected in the medical charts of all the patients involved in this observational study at the Bournigal Medical Center and the Punta Cana Medical Center.

## Authors contributions

All the authors involved actively participate in the definition of Grupo Rescue’s Covid-19 Management Guide, and are the directors of departments associated with the COVID-19 patient management program since the beginning of the pandemic. All of them have read and approved to publish this communication of the protocol and its preliminary results.

## Acknowledgments

The authors want to record the recognition of all health personnel involved in the successful management of COVID-19 cases, for their invaluable professional dedication and their permanent interest in collecting the data of the cases, in the computer matrix of the institutions involved. In addition, we want to thank the attitude of collaboration and loyalty of our patients, who have selected our emergency and Emergency Units to solve their health problems in this pandemic, and authorize through informed consents due to their inclusion in our management protocol.

